# Association analysis of mitochondrial DNA heteroplasmic variants: methods and application

**DOI:** 10.1101/2024.01.12.24301233

**Authors:** Xianbang Sun, Katia Bulekova, Jian Yang, Meng Lai, Achilleas N. Pitsillides, Xue Liu, Yuankai Zhang, Xiuqing Guo, Qian Yong, Laura M. Raffield, Jerome I. Rotter, Stephen S. Rich, Goncalo Abecasis, April P. Carson, Ramachandran S. Vasan, Joshua C. Bis, Bruce M. Psaty, Eric Boerwinkle, Annette L. Fitzpatrick, Claudia L. Satizabal, Dan E. Arking, Jun Ding, Daniel Levy, TOPMed mtDNA working group, Chunyu Liu

## Abstract

We rigorously assessed a comprehensive association testing framework for heteroplasmy, employing both simulated and real-world data. This framework employed a variant allele fraction (VAF) threshold and harnessed multiple gene-based tests for robust identification and association testing of heteroplasmy. Our simulation studies demonstrated that gene-based tests maintained an appropriate type I error rate at α=0.001. Notably, when 5% or more heteroplasmic variants within a target region were linked to an outcome, burden-extension tests (including the adaptive burden test, variable threshold burden test, and z-score weighting burden test) outperformed the sequence kernel association test (SKAT) and the original burden test. Applying this framework, we conducted association analyses on whole-blood derived heteroplasmy in 17,507 individuals of African and European ancestries (31% of African Ancestry, mean age of 62, with 58% women) with whole genome sequencing data. We performed both cohort- and ancestry-specific association analyses, followed by meta-analysis on bothpooled samples and within each ancestry group. Our results suggest that mtDNA-Enco ded genes/regions are likely to exhibit varying rates in somatic aging, with the notably strong associations observed between heteroplasmy in the *RNR1* and *RNR2* genes (*p*<0.001) and advance aging by the Original Burden test. In contrast, SKAT identified significant associations (*p*<0.001) between diabetes and the aggregated effects of heteroplasmy in several protein-coding genes. Further research is warranted to validate these findings. In summary, our proposed statistical framework represents a valuable tool for facilitating association testing of heteroplasmy with disease traits in large human populations.

## INTRODUCTION

Mitochondria are important organelles producing cellular energy through oxidative phosphorylation, calcium homeostasis, regulation of innate immunity, programmed cell death and stem cell regulation.(1) The maternally inherited mitochondrial genome is a circular molecule of double-stranded DNA (mtDNA). Human mtDNA consists of 16,569 base pairs and is essential for proper mitochondrial function. mtDNA encodes 22 tRNAs and 2 rRNAs, and 13 proteins that are involved in the energy production pathway (1) Hundreds to thousands of mtDNA molecules are present per human cell, depending on the cell’s energy requirement.(2) Heteroplasmy refers to a phenomenon where two or more alleles coexist at the same site in a mixture of mtDNA molecules within a cell or an individual.(3) Based on our previous studies(4) and other studies,(5-7) 98% of mtDNA heteroplasmic variants are rare, only present in one (i.e., singleton) or a few individuals. In addition, most heteroplasmic variants display low variant allele fractions (VAFs) in the general human population.(4-7) Nonetheless, the increase in both their number and VAFs of heteroplasmy during aging may contribute to age-related diseases, including cardiovascular disease and cancer.(2, 8)

With a greatly reduced cost in next generation sequencing technologies, hundreds of thousands of human genome samples, including mtDNA, have been sequenced. The availability of mtDNA sequences with high coverage (e.g., > 2000-fold) in large human populations(4) provides for the detection of rare, low-level heteroplasmic variants that are potentially associated with disease traits. The commonly used statistical methods to analyze rare variants in the nuclear genome, e.g., burden tests (9) and the sequence kernel association test (SKAT),(10) have not been evaluated for their performance in the context of ultra-rare variants such as mitochondrial heteroplasmy. In addition, there is no standard procedure or approach for the analysis of heteroplasmy. Therefore, it is important to develop a novel framework for testing the association of heteroplasmic variants with disease traits.

We propose a statistical framework for association analysis of heteroplasmic variants with a trait. This framework incorporates a pre-specified threshold for identifying true heteroplasmy and performs association analyses with a few methods, including the Original Burden test(9) and its extensions,(11, 12) and SKAT.(10) This framework also uses an aggregated Cauchy association test (ACAT-O)(13) and SKAT-optimal (SKAT-O)(14) to combine information from multiple gene-based methods applied in association analyses. Furthermore, this framework can easily incorporate different types of weights (e.g., the variant allele fraction and the predicted functional score). In this study, we evaluate the performance of these methods using simulated and real data to assess the association of heteroplasmy with continuous and binary traits.

## METHODS

### Definition of mitochondrial DNA sequence variants

Variant alleles are identified by comparing sequence reads in mtDNA to reference sequence, e.g., the revised Cambridge Reference Sequence (rCRS)(15) or Reconstructed Sapiens Reference Sequence (RSRS)(16). A variant allele fraction (VAF) is the proportion of the variant alleles over all sequence reads observed at a mtDNA site in an individual. To minimize false positive findings, a heteroplasmy is defined by a pre-specified threshold *τ* = (*τ*_1_, *τ*_2_). Let *VAF*_*ij*_ be the VAF of a variant at mtDNA site *j*^th^ in the *i*^th^ individual. Here *j* = 1, …, *m*, and *i* = 1, …, *n*. A site *j* is not considered as a variant if *VAF*_*ij*_ < *t*_1_ in individual *i*; it is considered as a heteroplasmy if *τ*_*1*_ ≤ *VAF*_*ij*_ ≤ *τ*_2_; and it is considered as a homoplasmy if *VAF*_*ij*_ > *r*_2_.

Let *G*_*ijt*_ be the coding of the heteroplasmy at the *j*^*th*^ site of *i*^*th*^ individual with a VAF threshold *r*. We consider two coding schemes for variants in association testing. First, we define a heteroplasmic variant by an indicator function in which the VAF of a heteroplasmy is not incorporated:

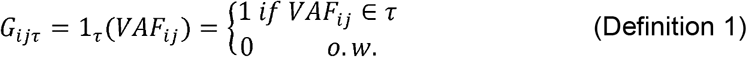

Second, we define a heteroplasmy by incorporating its VAF:

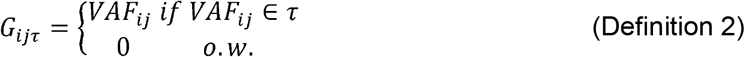

Because definition 2 results in distinct scales between heteroplasmic sites, we standardize the coding of each heteroplasmy, which is equivalent to the inverse variance weight using VAF.(10)

### A phenotype model

For subject *i*, let *y*_*i*_ denote a phenotype with mean *µ*_*i*_. Let *X*_*i*_ = (*X*_*i*1_,*…, X*_*iq*_)^*T*^ denote a vector of covariates, and let *G*_*iτ*_ = (*G*_*i*1*τ*_,*…, G*_*imr*_)^*T*^ be a vector of the coding of *m* mtDNA heteroplasmic sites in a region or gene. We consider a generalized linear mixed model (GLMM) framework to investigate the relationship between a set of mtDNA heteroplasmic variants in a region or gene and a phenotype.(17)

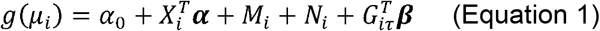

where *g*(*µ*_*i*_) = *µ*_*i*_ for a continuous trait and *g*(*µ*_*i*_) = *logit*(*µ*_*i*_) for a binary trait. To be more generalizable, we let *M*_*i*_ be a polygenic component of the mtDNA, and *N*_*i*_ be a polygenic component of the nuclear genome. In Equation 1, *α*_0_ is an intercept, ***α*** = *α*_1_,…, *α*_*q*_)^*T*^ is a column vector of the effects from covariates, and *β* = (*β*_*1*_,…, *β*_*m*_)^*T*^ is a column vector of the effects from a set of heteroplasmic variants. In this study, we only focus on mtDNA sequence variations, and therefore, we set *N*_*i*_ = 0. The testing of null hypothesis of no association between mtDNA sequence variations and a trait is equivalent to testing *H*_0_: ***β*** = (*β*_1_,…, *β*_*m*_)= **0**. The score statistic for mutation *j* is defined as

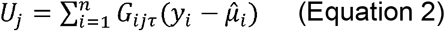

where 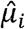 is the estimated mean of *y*_*i*_ under the null hypothesis (*H*_0_: *β* = 0) by fitting the null model 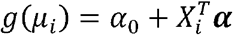

### Gene-based tests

#### The Original Burden and SKAT

In this study, we only considered heteroplasmic variants with population level frequency 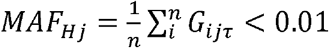 in association analyses. Here, *n* is the number of participants in the study and *MAF*_*Hj*_ refers to the minor allele frequency (MAF) of a heteroplasmic variant (H) *j*. The Original Burden test(9) (referred as Burden) and SKAT(10) are often used to aggregate the effects of rare variants in a genetic region in autosome. The corresponding test statistic for Burden is 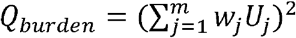. Under the null, *Q*_*burden*_ follows a chi-square distribution asymptotically with 1 degree of freedom. The SKAT method(10) uses variance component framework and the corresponding test statistic is 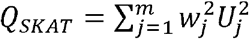. This test statistic follows a mixture of independent chi-square distributions asymptotically with 1 degree of freedom under the null. In both statistics for Burden and SKAT, *w*_*j*_ is a weight(18) that an investigator may choose for mutation *j*.

#### Extensions to the Original Burden test

Burden has larger power than SKAT when rare variants in a gene/region display the same effect direction in an association testing with a trait(14). Therefore, the adaptive burden test(11) (denoted as **Burden-A**) was proposed to improve power for Burden when variants have different effect directions in an association testing. The coding signs of rare variants are changed based on an arbitrary threshold of *p* value in the single variant model:

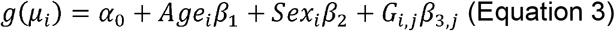

where g(.) is the identity function for a continuous trait and is the logistic function for a binary trait. The signs of the other variants remain the same. This leads to a new genetic dosage matrix *G*^*new*^ that contains variants with original signs and the ones after changing the signs. Then, the Original Burden test is performed with *G*^*new*^. By permuting the phenotype we generate an empirical null distribution of *p*_*new*_: {p^(*b)*^} with b=1,…, *B* based on a large number of *G*^*new*^ matrices. The empirical p value of the test is calculated as 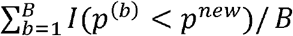. Here *p*^(*b)*^ or *p*^*new*^ is the *p* value based on the observed c matrix or the new matrix after switching the signs of certain heteroplasmic sites. We choose *B*=50000 for α level of 0.001 (**Supplemental Methods**).

Burden-A(11) uses the same weight by including both non-causal variants and causal variants in an association testing, which may lead to power loss. Sha and Zhang(12) proposed a z-score weighting approach (referred as **Burden-S**) to minimize this limitation: the z-score of j^th^ variant is calculated by 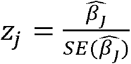 based on Equation 3, and the *z*_j_ score is used as weight in analysis with Burden-A. However, due to rareness of heteroplasmic variants, extreme z-scores may occur, which may lead to bias in association analyses. Therefore, we modify the z-score weights to have lower (z=-1.5) and upper (z=1.5) bounds. That is, we set *w*_j_ =1 if |*z*_*j*_| < z_0.05_ where z_0.05_ ≈ 1.65. If *z*_*j*_ ≥ *z*_0.05_, *w*_*j*_ is assigned to be *z*_*j*_ − *z*_0.05_ + 1, with an upper limit of 1.5. Similarly, if *z*_*j*_ ≤ −z_0.05_, *w*_*j*_ is assigned to be *z*_*j*_ + *z*_0.05_ − 1, with a lower limit of −1.5 (**Supplemental Methods**).

Another limitation of Burden-A is that the cutoff *p*_*C*_ based on marginal models is chosen arbitrarily. To overcome this, Sha and Zhang proposed the variable threshold approach(12) (**Burden-V**) that searches for an optimal cutoff in Burden-A based on Equation 3. This approach searches for all possible p values as the candidate thresholds,(12) which leads to an intensive computational burden. To decrease computational burden, we propose to use 15^th^, 30^th^, 50^th^, 70^th^ and 85^th^ percentiles (denoted by 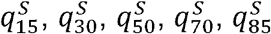 to be the thresholds with a continuous trait. We also perform the Original Burden test and obtain the p value *p*_0_. Based on these six p values 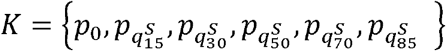, we define two test statistics: T_1_ = minK (**Burden-V1**) and T_2_ = ∑_*p*∈*K*_ *tan* ((0.5 − *p*) π) /|K| (**Burden-V2**). *T*_2_ is the test statistic of ACAT(13) (**Supplemental Methods**).

Because most of the heteroplasmic variants are singletons, a logistic regression with a binary trait leads to biased estimates and an extremely conservative p value *p*_3,*j*_ (>80% of the p values>0.9). Hence, for Burden-V1 and Burden-V2 with a binary trait, we modify the single mutation model, and fit a logistic regression under the null hypothesis of no genetic effect on the trait and obtain the residuals:

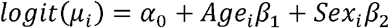

These residuals are rank-base inverse normalized. Then we regress the transformed residuals on each of the heteroplasmic variant to get the p value and beta coefficient. In addition, because a logistic regression with few rare mutations may lead to conservative results, we set the thresholds to be 50^th^, 70^th^ and 85^th^ percentiles.

#### The weights

The weights, *beta*(*MAF*, 1, 25), are used to put more weight on rarer variants in association analyses.(10) However, 98% of mtDNA heteroplasmic variants only present in one (i.e., singleton) or a few individuals.(4-7) Due to this extreme rareness, the beta(MAF, 1, 25) weights play a minimum role in the association analysis of rare heteroplasmic variants. For example, applying the *beta*(*MAF*, 1, 25) weights to analyzing a singleton heteroplasmy or a heteroplasmic variant of five individuals gives rise to almost identical weights (24.8 versus 24.0, respectively) in a cohort of 3,000 individuals (**Supplemental Results**). Simulation studies confirms the theoretical calculations (**Supplemental Figure 6**). Therefore, the *beta*(*MAF*, 1, 25) weights are not discussed in evaluating type I error and power.

### Combing multiple gene-based tests in a given gene/region

We adopt two omnibus tests (ACAT-O and SKAT-O) to combine information from association testing of heteroplasmic variants with Burden and SKAT. A generalized SKAT (SKAT-O)(14) is constructed as a linear combination of the Original Burden test and a SKAT(14). The test statistic is *Q*_*ρ*_ = *ρQ*_burden_ + (1 − *ρ*)*Q*_*SKAT*_, where *ρ*∈[0,1] is a weight for the Original Burden test. Under the null hypothesis, *Q*_*ρ*_ follows a mixture of independent chi-square distributions asymptotically with 1 degree of freedom. The SKAT-O can be constructed if we choose the minimum p value of the different choices of *ρ*,. The test statistic of SKAT-O is *Q*_*SKAT*-O_ = min {*pρ*1,…, p_*ρk*_}. The significance of *Q*_*SKAT*-O_ can be assessed by a one-dimensional numerical integration. In ACAT-O(13), the test statistic of a given region/gene is defined as 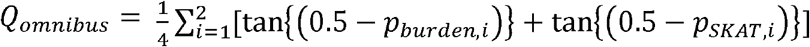. Here *p*_*burnet,i*_ and *p*_*SKAT,i*_ (*i*=1, 2) denote the p values from Burden test and the SKAT. Because this test statistic approximately follows a standard Cauchy distribution (13), the p value of the test statistic can be approximated by 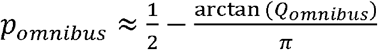.The ACAT-O method is computational efficient and it efficiently combines p values from individual tests of different methods when multiple weighting schemes are applied. In our study, to make it comparable to ACAT-O, we take *ρ*=0 and 1 to combine Burden and SKAT. That is, 0= *ρ*_1_ < *ρ*_2_ = 1. Based on previous studies, we use α=0.001 to control for multiple testing in association analyses across multiple genes/regions in simulation studies.(19)

### A simulation study

According to Equation 1, we simulated a continuous trait and a binary trait in response to heteroplasmic sites located in the mitochondrial Cytochrome b (MT-CYB) gene in European American participants (N=3,415) of Atherosclerosis Risk in Communities (ARIC) Study(20) (**Supplemental Information**). The CYB gene has a length of 1141 base pairs, and it is the fourth longest gene among the 13-mtDNA coding genes. Heteroplasmy was identified by a widely-used software package, MToolBox,(21) with WGS data by TOPMed Freeze 8, released in February 2019, GRCH38.(22) The rCRS(15) was used to identify heteroplasmic variants (**Supplemental Methods**). This gene contains 121 heteroplasmic sites in European American participants in ARIC. Of those, 68 are nonsynonymous and rare variants. The simulated traits were in response to these 68 nonsynonymous mutations (**Supplemental Table 1**) according to Equation 1. We simulated 50,000 replicates to evaluate the performance of the proposed methods with empirical type I error rate and power at □=0.001.

#### Type I error rate

To evaluate type I error rate, we simulated a phenotype, y, by Equation 1 under the null hypothesis: *y* = 0.08Age + Sex + *ε*, where *ε*∼N(0, 0.7). We applied a cutoff of 80% quantile to the simulated continuous phenotype to obtain a binary phenotype with 20% prevalence rate. The observed type I error rate is defined as the proportion of simulation replicates with p values ≤ 0.001 under the null. We evaluated type I error rate with a ratio of the observed type I error rate divided by 0.001. We used an arbitrary rule of thumb to evaluate if type I error rate is conservative or inflated based on the ratio: The type I error rate is conservative if the ratio is less than 0.4 and it is moderately conservative if the ratio is between 0.4 and 0.69. The type I error rate is appropriately controlled if the ratio is between 0.7 and 1.3. It is slightly inflated if the ratio is between 1.31 and 1.6 and inflated if the ratio is above 1.6.

#### Power estimation

To evaluate power, we simulated a continuous phenotype by the following model, a special case of Equation 1, with a genetic effect from sequence variations in the CYB gene: 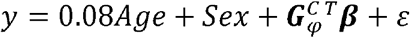, where 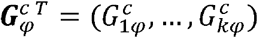 is a vector that includes of the coding for *k* randomly chosen causal heteroplasmic variants in the CYB gene. ***β***^T^ = (*β*_1_,…,*β*_*k*_) is a vector of fixed effects for the selected causal mutations. We also applied a cutoff of 80% quantile to the simulated continuous phenotype to obtain a binary phenotype. The effect size of heteroplasmic variant *j* is specified by 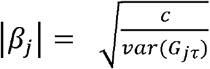, where *c* is a constant defined as 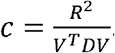. Here 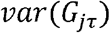 is the variance of the heteroplasmic variant *j, R*^2^ is the proportion of variance explained by all of the causal mutations, *D* is the correlation matrix between mutations, and *v* is a vector of the signs of ***β***. The proportion of variance (R^2^) explained by the causal heteroplasmic variants was set be 1% for the continuous phenotype, and 2% for the binary phenotype. The variance explained by age, sex and random error were around 99%.

#### Scenarios in simulation

In practice, it is possible that a proportion of heteroplasmic variants in a mtDNA gene posits effects on a phenotype while the rest of heteroplasmic variants in the same gene have no effects or display opposite effects on the phenotype. We considered that 5%, 25%, 50% and 80% of the nonsynonymous heteroplasmic variants in the CYB gene to be causal and 80% of these causal heteroplasmic variants to have the same effect direction with a phenotype. We further considered 50%, 100% of the causal mutations to have the same directionality in their associations with a phenotype. Therefore, we evaluated the power under 12 scenarios that vary the proportion of heteroplasmic variants to be causal and vary the proportion of the causal mutations to have the same directionality. To account for an inflated or conservative type I error rate, we estimate empirical power as the proportion of p values that is smaller than 0.1 quantile from all of the simulation replicates. We also present the power based on the nominal α level of 0.001. That is the proportion of simulation replicates with p values≤0.001.

### Application of the proposed framework in real data

#### Study participants

We applied the framework to analyze heteroplasmy with traits in five large cohorts, including ARIC(20), Framingham Heart Study (FHS)(23-25), Cardiovascular Health Study (CHS)(26), Jackson Heart Study (JHS)(27) and Multi-Ethnic Study of Atherosclerosis (MESA) (**Table 2, Supplemental information**).(28) These cohorts are prospective cohort studies that are aimed to investigate cardiovascular disease and its risk factors across different US populations. Participants in these five cohorts received whole genome sequencing (WGS) with an average coverage of 39-fold from the Trans-Omics for Precision Medicine (TOPMed) program, sponsored by the National Institutes of Health (NIH) National Heart, Lung and Blood Institute (NHLBI) (**Supplemental Methods**).(22) We excluded eleven duplicated participants between the cohorts (**Supplemental Methods**).

#### Identification of mtDNA heteroplasmy

Quality control of WGS sequencing was described previously(4) and was also briefly described in **Supplemental Methods**. We applied MToolBox(21) to all participating cohorts (WGS TOPMed Freeze 8, released in February 2019, GRCH38)(29) with rCRS.(15) We applied the 3%-97% of thresholds to identify heteroplasmy. The selection of the 3%-97% threshold with TOPMed WGS data and the detailed information for quality control of mtDNA sequence variations was described previously (**Supplemental Methods**).(4)

#### Traits in real data application

We applied our methods in association analyses of two pair of traits with heteroplasmy using gene-based tests. The first pair of variables included age and sex. Advancing age was known to be associated with a higher level of heteroplasmic burden. In contrast, inconsistent findings were reported in the association analysis of sex with heteroplasmic burden. We also included a pair of clinical traits, fasting blood glucose (FBG, mg/dL) and diabetes. Morning fasting blood glucose (mg/dL) was measured in each participating cohorts. In the analysis of FBG, we removed participants with measured FBG levels≥126 mg/dL or with diabetes treatment. Diabetes was defined as having a fasting blood glucose level of ≥126 mg/dL or currently receiving medications to lower blood glucose levels to treat diabetes.

#### Association analysis of heteroplasmy with traits

We applied two coding definitions to evaluate these gene-based tests and omnibus methods to analyze heteroplasmy in cross-sectional association analyses of two pair of traits (age and sex, and FBG and diabetes) with rare heteroplasmy (*MAF*_*Hj*_ < 0.01) in 16 genes/regions with the GLMM(17) framework. In association analyses, we used the heteroplasmic variants in sixteen genes/regions as the predictor variables while the two pair of traits as outcome variables. Covariates include batch variables for all models. In analysis of age as the outcome, we adjusted for sex; in analysis of sex as the outcome, we adjusted for age. In analyses of FBG and diabetes, age, sex, and body mass index were adjusted in addition to batch variables. We performed cohort-specific and ancestry-specific association analyses with the gene-based tests and omnibus tests (**Supplemental Methods**). Based on our simulation results, we applied the Burden-S method in addition to the Original Burden in association analyses of heteroplasmic variants with the traits. A previous study showed that year of blood draw was significantly associated with mtDNA copy number.(30) We tested if heteroplasmic burden was associated with year of blood draw in each cohort. We adjusted year of blood draw in cohort-specific association analyses if year of blood draw showed significant association with heteroplasmic burden in a cohort. White blood cell count/platelet showed associations with the total heteroplasmic burden,(4) therefore, we tested the association of heteroplasmy burden with blood cell count/platelet (**Supplemental Table 2**). We also performed sensitivity analyses with additionally adjusting for white blood cell count and differential count (the proportions of neutrophil, lymphocyte, monocyte, eosinophil, and basophil) and platelet variables. Because blood cell count/platelet variables were available in a subset of cohort/participants, the sensitivity analyses were performed in FHS (n=2551) and JHS (n=2737). We compared the beta estimates between the models with and without cell count/platelet variables using the same number of participants in these two cohorts.

Meta-analysis was used to combine results in participants of European ancestry (EA, n = 12,058, women 53.7%) and those of African ancestry (AA, n = 5460, women 60.8%) and in all participants of both ancestry. Ancestry-specific meta-analysis was performed in participants of European ancestry and African ancestry. Meta-analysis was also performed to combine results in all participants of both ancestries. We used two methods in conducting meta-analyses. We first combined the *p* values across cohorts by the Fisher’s method that does not employ weight in meta-analyses.(31) In addition, we performed a meta-analysis using the fixed-effects inverse variance method(32) to combine the summary statistics of the Original Burden test. Here, we hypothesized that there is only one true treatment effect for association of heteroplasmy with a trait between studies. We presented meta-analysis of all participants as the main result. For real data analyses, we used Bonferroni correction p<0.05/16∼0.003 for significance in association testing. All analyses in simulation and application used R software version 3.6.0.(33)

## RESULTS

We simulated a continuous trait and a binary trait based on heteroplasmic sites located in the mitochondrial Cytochrome b (MT-CYB) gene which is the forth longest gene of mtDNA (**Methods & Supplemental Table 1)**. Below we present results from simulation studies to evaluate type I error rate and power for several gene-based tests and the two omnibus tests. We also presented the findings from the application of these methods to real data in the five large cohorts with WGS.

### Empirical Type I Error Rate of simulation studies

We employ two coding definitions of heteroplasmy which are described thoroughly in methods section. By definition 1, for a continuous trait, type I error rate was appropriately controlled for Burden (ratio=0.88), Burden-A (ratio=1.22), Burden-S (ratio=1.06), and Burden-V1 (ratio=1.24); it was slightly inflated for the Burden-V2 (ratio=1.56) while moderately conservative for SKAT (ratio = 0.64) (**Table 1**). For the two omnibus tests, type I error rate was slightly conservative for SKAT-O (ratio=0.64) while appropriately controlled for ACAT-O (ratio=0.72). For a binary trait with a prevalence of 20%, type I error rate was appropriately controlled for Burden test (ratio =0.94), Burden-S (ratio=0.80) and SKAT (ratio=1.20), but slightly conservative for Burden-A (ratio=0.66) and conservative for Burden-V1 (ratio=0.32) and Burden-V2 (ratio=0.32). Both of the two omnibus tests have well-controlled type I error rates: SKAT-O (ratio=1.14) and ACAT-O (ratio=1.18) for a binary trait with a prevalence of 20% (**Table 1**).

**Table 1.**
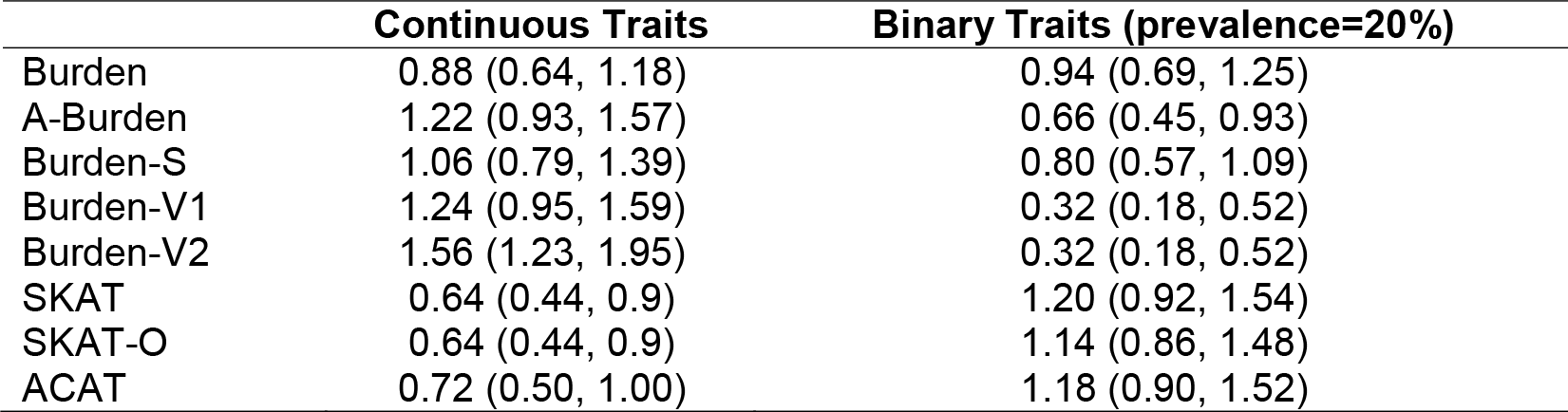
Gene-wide empirical type 1 error rates by coding definition 1 in. The number in each cell represents the ratio of type I error and expected significance level of 0.001. Burden, original burden test; Burden-A, adaptive burden test; Burden-S, z-score weighting burden test; Burden-V1, variable threshold burden test with minimum p value; Burden-V2, variable threshold burden test with ACAT p value combination method; SKAT, sequence kernel association test; SKAT-O, sequence kernel association test-optimal test; ACAT, aggregated Cauchy association test combining burden and SKAT. We simulated 50,000 replicates for evaluating type I error rate. We simulated a continuous variable and a binary variable in response to heteroplasmies located in the mitochondrial cytochrome b (MT-CYB) gene in European American participants (N=3,415) of Atherosclerosis Risk in Communities (ARIC) Study.

By definition 2, the gene-based tests and omnibus methods gave rise to similar type I error rates to their counterparts by definition 1 for a continuous trait (**Supplemental Table 3**). With a binary trait, the type I error rate was properly controlled for Burden (ratio=1.06), Burden-A (ratio=1.00) and Burden-S (ratio=0.98) while conservative for SKAT (ratio=0.3) and extremely conservative for Burden-V1 (ratio=0.002) and Burden-V2 (ratio=0.002). The two omnibus tests, SKAT-O (ratio=0.50) and ACAT-O (ratio=0.72), had moderately conservative type I error rates (**Supplemental Table 3**).

Of note, the weight, beta (MAF, 1, 25), which is widely used in gene-based association testing of nDNA of rare variants showed no effect in association testing of heteroplasmy (**Supplemental Results**) and therefore, this weight was not evaluated in the subsequent results. We use equal weights for each heteroplasmic variant at population level for all of our analysis.

### Empirical Statistical Power of simulation studies

#### Gene-based tests by definition 1

We estimated empirical power using the proportion of p values that is smaller than 0.1 quantile in simulation studies (**Figure 1, Supplemental Figure 1**) and using the fixed LJ = 0.001 (**Supplemental Figure 2-3**). For both continuous and binary traits, as expected, for all gene-based tests, power was improved when the proportion of causal variants (of all variants) increased and/or when the proportion of causal variants with the same effect direction increased for both definitions (**Figure 1, Supplemental Figure 1-3**).

**Figure 1.**
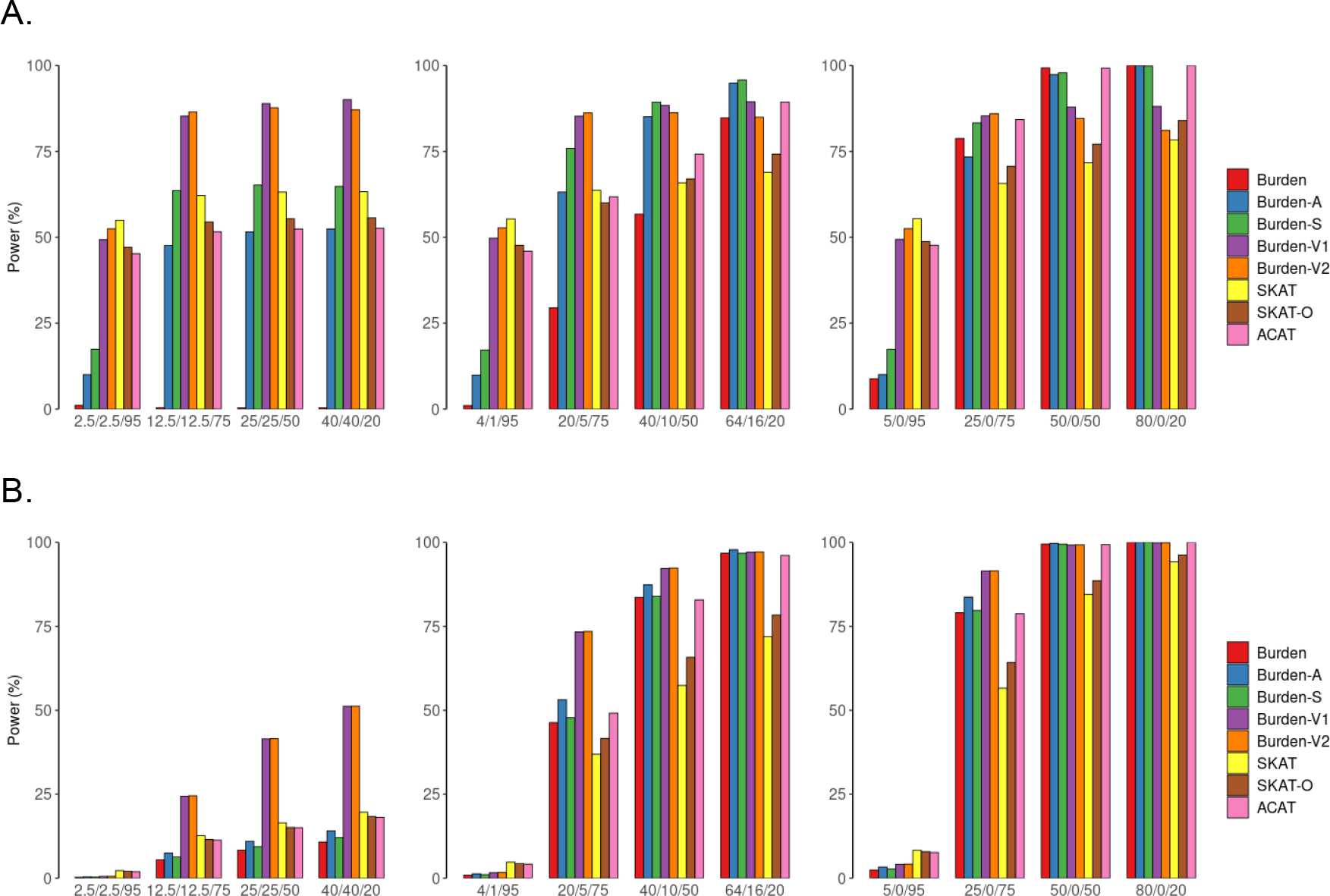
Simulation-based power comparisons of six gene-based tests and two omnibus tests with a continuous and a binary trait by coding definition 1 (adjusted for empirical type I error rate. Power estimation for a continuous trait (A) and a binary trait (B) at α=0.001. Heteroplasmic variants are defined by an indicator function (definition 1). In simulations, we consider 5%, 25%, 50% or 80% of the nonsynonymous heteroplasmies in CYB gene to be causal and consider that 50%, 80% and 100% of the causal heteroplasmic variants have effects with the same directionality. The variance that was explained by causal mutations was set to be 1% for the continuous trait and 2% for the binary trait. Burden, original burden test; Burden-A, adaptive burden test; Burden-S, z-score weighting burden test; Burden-V1, variable threshold burden test with minimum p value; Burden-V2, variable threshold burden test with ACAT p value combination method; SKAT, sequence kernel association test; SKAT-O, sequence kernel association test-optimal test; ACAT, aggregated Cauchy association test combining burden and SKAT. We simulated 50,000 replicates for evaluating power.

When 100% of heteroplasmic variants had the same effect direction, Burden and the burden extension methods displayed comparable power, adjusting for empirical alpha rate (**Figure 1, Supplemental Figure 1**). However, when any proportion of the causal variants displayed different effect directions, the burden extension methods, in general, outperformed the Burden method. Among these burden extension methods, Burden-V1 and Burden-V2 had comparable power under all scenarios; Burden-S, Burden-V1/V2 outperformed Burden-A when any proportion of heteroplasmic variants display different effect directions; and Burden-V1/V2 outperforms Burden S for most scenarios. For example, by definition 1, when 25% of heteroplasmic variants were causal, and 80% of these causal variants had the same effect direction, Burden had a low power (=0.29) while Burden-A (=0.63), Burden-S (=0.76), Burden-V1/V2 (=0.85) had much higher power (**Figure 1**).

For a continuous trait, if other conditions were held constant, SKAT outperformed all burden methods if 5% or less of heteroplasmic variants were causal in a region (**Figure 1**). Of note, the power was also low (<0.6) for SKAT if < 25% of heteroplasmic variants was causal. If the proportion of causal heteroplasmic variants increased to 25% or higher, all burden methods displayed comparable or higher power than SKAT. For example, when 50% of the heteroplasmic variants were causal and 50% of the causal variants had the same effect direction, SKAT had a power of 0.63, Burden-S had a power of 0.65, and Burden-V1/V2 had a power of 0.89.

For a binary trait, most burden tests had comparable or higher power than SKAT when the proportion of causal heteroplasmic variants was 25% or higher (**Figure 1**), regardless of the effect direction. For example, Burden-V1 exhibited 156% greater power than the SKAT (0.41 versus 0.16) when 50% of the heteroplasmic variants were causal and 50% of these causal heteroplasmic variants had the same effect directionality. When only 5% of the heteroplasmic variants were causal and 50% of them had the same effect direction, neither Burden-V1 nor SKAT had power (0.002 versus 0.003).

#### Two omnibus tests by definition 1

SKAT-O test had comparable power to SKAT under all scenarios. When other conditions were held constant, ACAT-O had a similar power to the more powerful gene-based test (i.e., a SKAT or Burden depending on the different scenarios), and therefore, ACAT-O was more powerful than SKAT-O when the real disease model was unknown.SKAT-O and ACAT-O displayed comparable power when 50% of the causal mutations had the opposite effect direction. However, ACAT-O was more powerful than SKAT-O if 80% or 100% of the causal mutations had the same effect direction (**Figure 1 & Supplemental Figure 2**).

#### Definition 2

In general, we observed consistent results by definition 2 compared to those by definition 1. In brief, by definition 2, for both continuous and binary traits, SKAT outperformed all Burden tests when 5% of heteroplasmic variants were causal and other conditions were fixed. For example, when 5% of the heteroplasmic variants were causal and 80% of these causal mutations had the same effect direction, the power of SKAT and Burden-V1 are 0.91 and 0.53, respectively, with a continuous trait. These two methods had comparable power if 25% or more heteroplasmic variants were causal. For example, these two methods had power of 0.92 when the proportion of causal heteroplasmic variants increased to 50% given other conditions were fixed. For the omnibus tests, SKAT-O had a great power loss for both continuous and binary traits with definition 2. For example, when 25% of the mutations were causal and 80% of the causal mutations had the same effect direction SKAT-O had power of 0.15 while ACAT-O had power of 0.56 for a binary trait (**Supplemental Figures 2, 3**).

### Application to real data

We identified heteroplasmic variants and performed quality control procedures in five TOPMed cohorts containing middle-aged and older participants [5456 African Americans (AA, mean age 59, women 61%) and 12,051 European Americans (EA, mean age 63, women 56%] (**Table 2, Supplemental Methods, Supplemental Table 4, Supplemental Figure 4**). Meta-analysis was used to combine results in participants of American Whites (EA, n = 12,058, women 53.7%) and those of African Americans (AA, n = 5460, women 60.8%) separately, and in all participants of both races. We reported meta-analysis results in all participants as the primary findings and compared findings between American Whites and African Americans.

**Table 2.** Participant characteristics in the five population level cohorts with whole genome sequencing.

| Cohort | Sample size | Age, mean (±SD) | Female, n (%) | FBG (±SD) | Diabetes, n (%) | n of heteroplasmy |
| --- | --- | --- | --- | --- | --- | --- |
| African American (n = 5456, women 60.8%) |  |  |  |  |  |  |
| ARIC | 241 | 58.4 (6.3) | 144 (59.8) | 100.2 (9.8) | 54 (25.5) | 162 |
| CHS | 705 | 73.8 (5.6) | 445 (63.1) | 97.3 (11.3) | 158 (23.8) | 673 |
| JHS | 3404 | 55.7 (12.8) | 2140 (62.9) | 90.5 (8.9) | 469 (21.3) | 1590 |
| MESA | 1106 | 60.9 (9.6) | 588 (53.1) | 90.1 (10.7) | 174 (15.9) | 968 |
| European American (n = 12,051, women 53.7%) |  |  |  |  |  |  |
| ARIC | 3415 | 58.2 (5.9) | 1734 (50.9) | 101.3 (9.6) | 368 (11.1) | 1501 |
| CHS | 2788 | 74.2 (5.7) | 1594 (57.2) | 98.6 (9.8) | 352 (13.1) | 1859 |
| FHS | 3992 | 59.9 (15.7) | 2190 (54.9) | 97.3 (10.2) | 175 (10.5) | 2158 |
| MESA | 1856 | 61.6 (9.8) | 952 (51.1) | 87.1 (9.6) | 95 (5.1) | 1236 |

ARIC, Atherosclerosis Risk in Communities Study; CHS, Cardiovascular Health Study; FHS, Framingham Heart Study; JHS, Jackson Heart Study; MESA, Multi-Ethnic Study of Atherosclerosis. SD, standard deviation

#### Association of heteroplasmy with age and sex

Two definitions of heteroplasmy coding tended to yield consistent p-values across methods in association testing with age in meta-analyses of all participants (**Supplemental Tables 5-8**) and in ancestry-specific meta-analyses (**Supplemental Tables 9-16**). In meta-analysis of all participants by the Fisher’s method, *RNR*1, *RNR*2, *CO*1, *CO*2, and *ND*4 showed significant associations with age (*p* < 0.001) using either definition 1 or definition 2 by multiple methods (**Table 3, Supplemental Tables 5-6**). Using the fixed effect inverse variance method of the Original Burden test, *D-loop, RNR1, RNR2, CO1*, CO3, and *ND4, ND5, CYB* showed significant associations with age (*p* < 0.001) (**Figure 2, Supplemental Tables 7-8**). Using *RNR1* as an example, an increase by one heteroplasmy (definition 1) in this gene was significantly associated with 1.09 years of older age (*p* = 4.9E-7) (**Supplemental Table 7**). In addition, an increase by 1 SD increase in heteroplasmy VAF (definition 2) in *RNR*1 was significantly associated with 0.036 years of older age (*p* = 5.5E-11) (**Supplemental Table 8**). Sensitivity analysis showed that association strength remained consistent after adjusting for white blood cell count, component counts and platelet counts (**Supplemental Figure 5**).

**Table 3.** Genes showing significant associations with age in meta-analysis using the Fisher’s method in pooled participants.

| mtDNA region | P values |  |  |  |  |  |  |  |
| --- | --- | --- | --- | --- | --- | --- | --- | --- |
|  | Burden | Burden-A | Burden-S | Burden-V1 | Burden-V2 | SKAT | SKAT-O | ACAT |
| Definition 1 |  |  |  |  |  |  |  |  |
| MT-*RNR1* | 1.1E-08 | 1.3E-08 | 4.2E-08 | 0.00012 | 0.0038 | 0.0072 | 6.2E-05 | 3.0E-08 |
| MT-*RNR2* | 2.5E-10 | 2.2E-10 | 2.0E-09 | 5.0E-04 | 5.9E-08 | 0.0073 | 1.6E-06 | 1.1E-08 |
| MT-*CO1* | 7.2E-05 | 1.5E-05 | 2.5E-05 | 0.062 | 0.00061 | 0.071 | 0.00037 | 0.00034 |
| Definition 2 |  |  |  |  |  |  |  |  |
| MT-*RNR1* | 5.5E-12 | 1.2E-08 | 5.0E-08 | 4.0E-05 | 1.5E-06 | 0.0014 | 5.3E-05 | 6.3E-11 |
| MT-*RNR2* | 6.9E-12 | 2.2E-10 | 2.0E-09 | 0.00039 | 4.2E-08 | 0.014 | 0.00082 | 2.4E-10 |
| MT-*CO1* | 1.9E-06 | 3.2E-06 | 4.9E-06 | 0.049 | 5.8E-05 | 0.05 | 0.0025 | 2.1E-05 |
| MT-*CO2* | 0.011 | 0.025 | 0.021 | 0.053 | 0.042 | 0.0061 | 0.00061 | 0.0014 |
| MT-*ND4* | 0.0046 | 0.0049 | 0.0013 | 0.086 | 0.09 | 0.18 | 0.23 | 0.015 |

Cohort-specific association analysis was performed between heteroplasmic variants and age using gene-based tests and omnibus tests. Fisher's method was used to combine p values from individual cohort/ancestry (i.e., meta-analysis) from all participants. This table include genes that yielded $p \leq 0.001$ in any gene based tests after meta-analysis. Burden, original burden test; Burden-A, adaptive burden test; Burden-S, z-score weighting burden test; Burden-V1, variable threshold burden test with minimum p value; Burden-V2, variable threshold burden test with ACAT p value combination method; SKAT, sequence kernel association test; SKAT-O, sequence kernel association test-optimal test; ACAT, aggregated Cauchy association test combining burden and SKAT.

**Figure 2.**
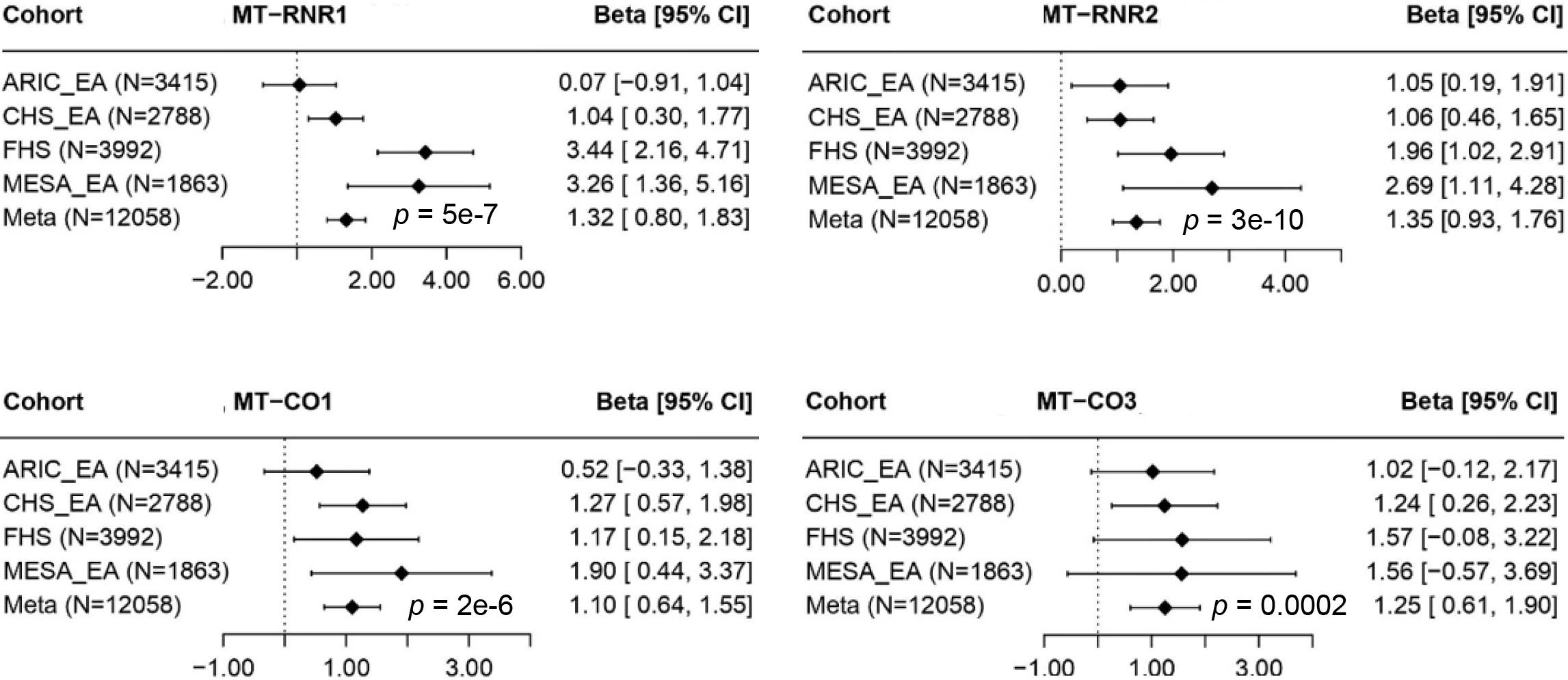
Examples of significant age-associated genes in European Americans (EA). ARIC, Atherosclerosis Risk in Communities (ARIC) Study; FHS, Framingham Heart Study, CHS, Cardiovascular Health Study; MESA, Multi-Ethnic Study of Atherosclerosis.

In meta-analysis of African Americans by Fisher’s method for an association with age, *RNR2* was the only gene showing significant association (*p* < 0.001) by multiple burden tests but not by SKAT with the two heteroplasmy definitions (**Supplemental Tables 9-10)**. The fixed effect inverse variance method also yielded significant findings between the *RNR2* gene and age with the two definitions (**Supplemental Tables 11-12**). In meta-analysis of EA participants, multiple genes, including *RNR1, RNR2, CO1, CO2*, and *ND4*, showed significant associations with age (*p* < 0.001) by multiple tests (**Supplemental Tables 13-16**). Due to the dominant sample size, the results in meta-analysis of EA participants largely represented the findings in meta-analysis of all participants (**Supplemental Tables 5-8**).

Unlike the findings in association and meta-analysis with age, heteroplasmy in most of mtDNA genes showed no association with sex (**Supplemental Tables 17-28**). *ND5* was the only gene associated with sex (*p* < 0.001) in meta-analysis of all participants and AA participants using the Burden-V2 method by definition 2 (**Supplemental Tables 18 & 22**). In meta-analysis with participants of EA, no genes showed significant associations with sex (all *p* > 0.001) (**Supplemental Tables 25-28**).

#### Association of heteroplasmy with fasting blood glucose and diabetes

Heteroplasmy showed no association (*p*<0.001) with FBG in any of the sixteen genes with any of the gene-based or omnibus tests by the Fisher’s method or fixed effect method in meta-analysis of all participants or race-specific samples (**Supplemental Tables 29-40**). In association analysis with diabetes (**Supplemental Tables 41-52**), the heteroplasmy in three genes, *CO3* (p=0.00047), *ND1* (p=4.0E-04), *and ND6* (p=7.9E-06) displayed significant associations (*p* < 0.001) by SKAT using definition 1 and the Fisher’s method in meta-analysis of all participants (**Table 4 & Supplemental Table 41**). However, the Original Burden and Burden-S methods did not give rise to any significant associations between heteroplasmy and diabetes in meta-analysis of all participants or race-specific samples (**Supplemental Table 41-52**).

**Table 4.** Genes showing significant associations with diabetes in meta-analysis using the Fisher’s method in pooled participants by definition 1.

| Gene | Burden | Burden-S | SKAT | SKAT-O | ACAT |
| --- | --- | --- | --- | --- | --- |
| MT-ND1 | 0.31 | 0.32 | 0.00047 | 0.0018 | 0.0085 |
| MT-CO3 | 0.11 | 0.11 | 4.0E-04 | 0.0012 | 0.0017 |
| MT-ND5 | 0.15 | 0.25 | 0.0022 | 0.0035 | 0.0033 |
| MT-ND6 | 0.17 | 0.071 | 7.9E-06 | 2.0E-05 | 5.0E-05 |

Cohort-specific association analysis was performed between heteroplasmic variants and diabetes using gene-based tests and omnibus tests. Fisher's method was used to combine p values from individual cohort/ancestry (i.e., meta-analysis) from all participants. This table includes genes that yielded $p \leq 0.003$ in any gene based tests after meta-analysis. Burden, original burden test; Burden-A, adaptive burden test; Burden-S, z-score weighting burden test; SKAT, sequence kernel association test; SKAT-O, sequence kernel association test-optimal test; ACAT, aggregated Cauchy association test combining burden and SKAT.

## Discussion

We proposed a framework that incorporates a pre-specified threshold for identifying true heteroplasmic variants and several gene-based tests to perform association analyses between heteroplasmic variants and a trait. We used simulation studies to evaluate the proposed framework in association analyses of mtDNA heteroplasmic variants and applied this framework to analyze age and sex with rare heteroplasmic variants in five large TOPMed cohorts with WGS.

### Simulations studies

The proposed framework incorporates several gene-based methods and omnibus test to provide a comprehensive evaluation of trait-heteroplasmy association. The burden-extension tests outperformed the SKAT method for all simulation scenarios except for extreme unfavorable situations in which a very small proportion (≤5%) of the heteroplasmic variants were causal and/or half of these causal variants display opposite directions. Under such unfavorable situations, the Original Burden had almost no power while the burden-extension tests had comparable power to SKAT. The Original Burden showed comparable power to burden extension methods only when ∼100% of heteroplasmic variants showed consistent effect direction. Of the two omnibus tests, ACAT-O easily combines a large number of test p-values and it was more powerful than SKAT-O for most situations when combining SKAT and the Original Burden test.

It is worth noting that the widely used weights, i.e., beta (MAF, 1, 25), in association testing of nDNA rare variants showed no effects in testing of rare heteroplasmic variants, owing to the extreme rareness of heteroplasmic variants in human population. While these methods outperformed the Original Burden test, the burden-extension tests provided only p values without computing effect size for a gene. The burden-extension tests use permutation to derive p values, which is computationally extensive. These extension methods are challenging to analyzing a large number genes in nuclear DNA while they are feasible in analyzing a small number of genes in mtDNA. The utilization of multiple burden methods offers valuable insights into the proportion of heteroplasmic variants linked to the trait and their directional effects in gene-based tests. We are currently extending the framework to identify these trait-associated heteroplasmic variants and classify the trait-associated heteroplasmic variants into distinct groups with different effect directions.

### Association studies in real data

It is known that heteroplasmy burden is increase with advancing aging.(4, 34) However, few studies have investigated rare heteroplasmic mutations specific genes with aging. The Original Burden test uncovered significant associations of advancing age with the number and alternative allele fractions of heteroplasmic variants in both non-protein-coding regions and protein-coding genes. These findings indicate that the 16 mtDNA-encoded genes/regions are likely to vary their rates in somatic aging, with the most pronounced associations observed in the *RNR1* and *RNR2* genes that encode a 12S rRNA and 16S rRNA, respectively. The two rRNA molecules are part of the machinery for the synthesis of 13 mtDNA-encoded polypeptides that are essential components of the mitochondrial oxidative phosphorylation (OXPHOS) pathway.(35) Despite their key roles in mitochondrial biogenesis, these two genes have been studied in far less detail than protein-coding genes in mtDNA with regard to their associations with disease traits. Mutations in *RNR1* were found to cause hearing loss.(36, 37) More recently, a small open reading frame within *RNR2* that encodes the humanin polypeptide has been the target of Alzheimer’s disease research.(38, 39) Given that aging is the leading cause for Alzheimer’s disease, heteroplasmic variants in these two genes merit further investigations for their relationships with Alzheimer’s disease and other age-related diseases. Additional significant genes associated with advancing age are three protein-coding genes in *CO1, CO2* and *ND4*. mtDNA encodes the three largest subunits of the cytochrome c oxidase (COX) genes (I, II, and III) for complex IV of the terminal OXPHOS respiratory chain, which is crucial for aerobic metabolism.(40) Maternally inherited mutations in the CO subunits are associated with many severe, inherited mitochondrial diseases.(41-43) The nicotinamide adenine dinucleotide (NADH)-ubiquinone oxidoreductase 4 (*ND4*) gene is one of the seven genes encoded by mtDNA for complex I.(44) Mutations in the *ND4* gene has been linked to optic nerve atrophies(45) and multiple sclerosis(46).

In contrast to the Original Burden test that identified the most significant heteroplasmy-age associations, the SKAT test identified the most significant associations between diabetes and the aggregation effects of heteroplasmies in only protein-coding genes using definition 1. Given the properties of the original burden test and SKAT test, it is reasonable to speculate that the heteroplasmic variants in several protein-coding genes are likely to exhibit different magnitudes and/or opposite association directions with diabetes. The maternally inherited insulin-dependent(47-52) and noninsulin-dependent(53-56) diabetes have been linked to point mutations in the mtDNA coded tRNA genes(47-51) and several ND genes (*ND1, ND2*, and *ND6*) in complex I of OXPHOS. However, the links between rare heteroplasmic mutations and diabetes have not been documented thus far. Our analyses revealed that the aggregation effects of rare heteroplasmic variants in *CO3, ND1, ND5*, and *ND6* were significantly associated with higher chance of diabetes. The *CO3* gene produces a protein that is a member of the cytochrome c oxidase subunit 3 family. This protein is located on the inner mitochondrial membrane. As pointed out in the preceding paragraph, *CO3* is located in the terminal Complex IV of the OXPHOS respiratory chain for aerobic metabolism. The ND complex is the first and the largest complex of the electron transport chain.(44) Complex 1 oxidizes nicotinamide adenine dinucleotide (NADH) to generate electrons from NADH to coenzyme Q10 (CoQ10) and translocates protons across the inner mitochondrial membrane for energy metabolism.(44) Point mutations in the three CO genes of complex IV have not be reported with insulin-dependent or noninsulin-dependent diabetes.

In summary, the proposed framework provides a comprehensive evaluation of trait-heteroplasmy association. Using this framework, we found that heteroplasmic variants are not likely to differ between men and women. We found that somatic aging occurs unevenly across mtDNA regions. We also found that aggregation effects of rare heteroplasmic variants in a few gens were associated with diabetes. These findings merits further investigation in independent cohorts. This framework will facilitate association analyses of heteroplasmic variants with complex, age-related traits in large population data with WGS.

## Data Availability

All data produced are available online at
https://www.ncbi.nlm.nih.gov/gap/

https://www.ncbi.nlm.nih.gov/gap/

## Acknowledgments

We included detailed acknowledgment for each cohort in Supplemental Materials. We thank the staff and participants of the ARIC, CHD, FHS, JHS, and MESA cohorts for phenotype data collections and providing biological samples and data for TOPMed. Whole genome sequencing (WGS) for the Trans-Omics in Precision Medicine (TOPMed) program was supported by the National Heart, Lung and Blood Institute (NHLBI). Centralized read mapping and genotype calling, along with variant quality metrics and filtering were provided by the TOPMed Informatics Research Center (R01HL-117626-02S1; contract HHSN268201800002I). Phenotype harmonization, data management, sample-identity QC, and general study coordination were provided by the TOPMed Data Coordinating Center (R01HL-120393-02S1; contract HHSN268201800001I). Method development and statistical analysis was supported by R21HL144877 (X.S., K.B., A.P., and C.L.), R01AG059727 (X.L, C.L., and C.S.) and R01HL15569 (M.L.). The views expressed in this manuscript are. those of the authors and do not necessarily represent the views of the National Heart, Lung, and Blood Institute; the National Institutes of Health; or the U.S. Department of Health and Human Services.

## Author contributions

**Data preparation**, X.S., M.L., A.P., X.L., T.B., X.G., L.M.R. X.G., Y.Z., G.A., J.C.B.; **mtDNA heteroplasmy identification**: X.S., K.B., M.L., A.P. Q.Y. J.D.; **Statistical analyses**: X.S., K.B., M.L., A.P.; **Manuscript preparation and revision**: X.L., C.L. Y.Z., J.C.B., A.L.F., S.R.H.; D.A., D.L.; **Funding support**: C.L., C.L.S., J.I.R., S.S.R., A.C., M.F., B.M.P., E.B., J.G.W.

## REFERENCES

1 Voet, D., Voet, J.G. and Pratt, C.W. (2006) Fundamentals of biochemistry : life at the molecular level. Wiley, Hoboken, N.J.

2 Wallace, D.C. (2013) A mitochondrial bioenergetic etiology of disease. J Clin Invest, 123, 1405–1412.

3 Wallace, D.C. (2015) Mitochondrial DNA variation in human radiation and disease. Cell, 163, 33–38.

4 Liu, C., Fetterman, J.L., Qian, Y., Sun, X., Blackwell, T.W., Pitsillides, A., Cade, B.E., Wang, H., Raffield, L.M., Lange, L.A. et al. (2021) Presence and transmission of mitochondrial heteroplasmic mutations in human populations of European and African ancestry. Mitochondrion, 60, 33–42.

5 Ding, J., Sidore, C., Butler, T.J., Wing, M.K., Qian, Y., Meirelles, O., Busonero, F., Tsoi, L.C., Maschio, A., Angius, A. et al. (2015) Assessing Mitochondrial DNA Variation and Copy Number in Lymphocytes of ∼2,000 Sardinians Using Tailored Sequencing Analysis Tools. PLoS Genet, 11, e1005306.

6 Liu, C., Fetterman, J.L., Liu, P., Luo, Y., Larson, M.G., Vasan, R.S., Zhu, J. and Levy, D. (2018) Deep sequencing of the mitochondrial genome reveals common heteroplasmic sites in NADH dehydrogenase genes. Hum Genet, 137, 203–213.

7 Ye, K., Lu, J., Ma, F., Keinan, A. and Gu, Z. (2014) Extensive pathogenicity of mitochondrial heteroplasmy in healthy human individuals. Proc Natl Acad Sci U S A, 111, 10654–10659.

8 Stewart, J.B. and Chinnery, P.F. (2015) The dynamics of mitochondrial DNA heteroplasmy: implications for human health and disease. Nat Rev Genet, 16, 530–542.

9 Li, B. and Leal, S.M. (2008) Methods for detecting associations with rare variants for common diseases: application to analysis of sequence data. Am J Hum Genet, 83, 311–321.

10 Wu, M.C., Lee, S., Cai, T., Li, Y., Boehnke, M. and Lin, X. (2011) Rare-variant association testing for sequencing data with the sequence kernel association test. Am J Hum Genet, 89, 82–93.

11 Han, F. and Pan, W. (2010) A data-adaptive sum test for disease association with multiple common or rare variants. Hum Hered, 70, 42–54.

12 Sha, Q., Wang, S. and Zhang, S. (2013) Adaptive clustering and adaptive weighting methods to detect disease associated rare variants. Eur J Hum Genet, 21, 332–337.

13 Liu, Y., Chen, S., Li, Z., Morrison, A.C., Boerwinkle, E. and Lin, X. (2019) ACAT: A Fast and Powerful p Value Combination Method for Rare-Variant Analysis in Sequencing Studies. Am J Hum Genet, 104, 410–421.

14 Lee, S., Wu, M.C. and Lin, X. (2012) Optimal tests for rare variant effects in sequencing association studies. Biostatistics, 13, 762–775.

15 Andrews, R.M., Kubacka, I., Chinnery, P.F., Lightowlers, R.N., Turnbull, D.M. and Howell, N. (1999) Reanalysis and revision of the Cambridge reference sequence for human mitochondrial DNA. Nat Genet, 23, 147.

16 Behar, D.M., van Oven, M., Rosset, S., Metspalu, M., Loogvali, E.L., Silva, N.M., Kivisild, T., Torroni, A. and Villems, R. (2012) A “Copernican” reassessment of the human mitochondrial DNA tree from its root. Am J Hum Genet, 90, 675–684.

17 Chen, H., Huffman, J.E., Brody, J.A., Wang, C., Lee, S., Li, Z., Gogarten, S.M., Sofer, T., Bielak, L.F., Bis, J.C. et al. (2019) Efficient Variant Set Mixed Model Association Tests for Continuous and Binary Traits in Large-Scale Whole-Genome Sequencing Studies. Am J Hum Genet, 104, 260–274.

18 Lee, S., Abecasis, G.R., Boehnke, M. and Lin, X. (2014) Rare-variant association analysis: study designs and statistical tests. Am J Hum Genet, 95, 5–23.

19 Kraja, A.T., Liu, C., Fetterman, J.L., Graff, M., Have, C.T., Gu, C., Yanek, L.R., Feitosa, M.F., Arking, D.E., Chasman, D.I. et al. (2019) Associations of Mitochondrial and Nuclear Mitochondrial Variants and Genes with Seven Metabolic Traits. Am J Hum Genet, 104, 112–138.

20 (1989) The Atherosclerosis Risk in Communities (ARIC) Study: design and objectives. The ARIC investigators. Am J Epidemiol, 129, 687–702.

21 Calabrese, C., Simone, D., Diroma, M.A., Santorsola, M., Gutta, C., Gasparre, G., Picardi, E., Pesole, G. and Attimonelli, M. (2014) MToolBox: a highly automated pipeline for heteroplasmy annotation and prioritization analysis of human mitochondrial variants in high-throughput sequencing. Bioinformatics, 30, 3115–3117.

22 Taliun, D., Harris, D.N., Kessler, M.D., Carlson, J., Szpiech, Z.A., Torres, R., Taliun, S.A.G., Corvelo, A., Gogarten, S.M., Kang, H.M. et al. (2021) Sequencing of 53,831 diverse genomes from the NHLBI TOPMed Program. Nature, 590, 290–299.

23 Dawber, T.R., Meadors, G.F. and Moore, F.E., Jr. (1951) Epidemiological approaches to heart disease: the Framingham Study. Am J Public Health Nations Health, 41, 279–281.

24 Feinleib, M., Kannel, W.B., Garrison, R.J., McNamara, P.M. and Castelli, W.P. (1975) The Framingham Offspring Study. Design and preliminary data. Prev Med, 4, 518–525.

25 Splansky, G.L., Corey, D., Yang, Q., Atwood, L.D., Cupples, L.A., Benjamin, E.J., D’Agostino, R.B., Sr. Fox, C.S., Larson, M.G., Murabito, J.M. et al. (2007) The Third Generation Cohort of the National Heart, Lung, and Blood Institute’s Framingham Heart Study: design, recruitment, and initial examination. Am J Epidemiol, 165, 1328–1335.

26 Fried, L.P., Borhani, N.O., Enright, P., Furberg, C.D., Gardin, J.M., Kronmal, R.A., Kuller, L.H., Manolio, T.A., Mittelmark, M.B., Newman, A. et al. (1991) The Cardiovascular Health Study: design and rationale. Ann Epidemiol, 1, 263–276.

27 Wilson, J.G., Rotimi, C.N., Ekunwe, L., Royal, C.D., Crump, M.E., Wyatt, S.B., Steffes, M.W., Adeyemo, A., Zhou, J., Taylor, H.A., Jr. et al. (2005) Study design for genetic analysis in the Jackson Heart Study. Ethn Dis, 15, S6–30-37.

28 Bild, D.E., Bluemke, D.A., Burke, G.L., Detrano, R., Diez Roux, A.V., Folsom, A.R., Greenland, P., Jacob, D.R., Jr., Kronmal, R., Liu, K. et al. (2002) Multi-Ethnic Study of Atherosclerosis: objectives and design. Am J Epidemiol, 156, 871–881.

29 (2021), in press.

30 Liu, X., Longchamps, R.J., Wiggins, K.L., Raffield, L.M., Bielak, L.F., Zhao, W., Pitsillides, A., Blackwell, T.W., Yao, J., Guo, X. et al. (2021) Association of mitochondrial DNA copy number with cardiometabolic diseases. Cell Genomics, 1.

31 Fisher, R.A. (1925) Statistical methods for research workers. Oliver and Boyd, Edinburgh, London,.

32 Borenstein, M., Hedges, L.V., Higgins, J.P. and Rothstein, H.R. (2010) A basic introduction to fixed-effect and random-effects models for meta-analysis. Res Synth Methods, 1, 97–111.

33 R Core Team. (2019) R: A language and environment for statistical computing. Journal, in press.

34 Sondheimer, N., Glatz, C.E., Tirone, J.E., Deardorff, M.A., Krieger, A.M. and Hakonarson, H. (2011) Neutral mitochondrial heteroplasmy and the influence of aging. Hum Mol Genet, 20, 1653–1659.

35 Scheffler, I.E. (2008) Mitochondria. Wiley-Liss, Hoboken, N.J.

36 Li, R., Greinwald, J.H., Jr., Yang, L., Choo, D.I., Wenstrup, R.J. and Guan, M.X. (2004) Molecular analysis of the mitochondrial 12S rRNA and tRNASer(UCN) genes in paediatric subjects with non-syndromic hearing loss. J Med Genet, 41, 615–620.

37 Yao, Y.G., Salas, A., Bravi, C.M. and Bandelt, H.J. (2006) A reappraisal of complete mtDNA variation in East Asian families with hearing impairment. Hum Genet, 119, 505–515.

38 Yen, K., Mehta, H.H., Kim, S.J., Lue, Y., Hoang, J., Guerrero, N., Port, J., Bi, Q., Navarrete, G., Brandhorst, S. et al. (2020) The mitochondrial derived peptide humanin is a regulator of lifespan and healthspan. Aging (Albany NY), 12, 11185–11199.

39 Tajima, H., Niikura, T., Hashimoto, Y., Ito, Y., Kita, Y., Terashita, K., Yamazaki, K., Koto, A., Aiso, S. and Nishimoto, I. (2002) Evidence for in vivo production of Humanin peptide, a neuroprotective factor against Alzheimer’s disease-related insults. Neurosci Lett, 324, 227–231.

40 Fontanesi, F., Soto, I.C. and Barrientos, A. (2008) Cytochrome c oxidase biogenesis: new levels of regulation. IUBMB Life, 60, 557–568.

41 Brown, M.D., Yang, C.C., Trounce, I., Torroni, A., Lott, M.T. and Wallace, D.C. (1992) A mitochondrial DNA variant, identified in Leber hereditary optic neuropathy patients, which extends the amino acid sequence of cytochrome c oxidase subunit I. Am J Hum Genet, 51, 378–385.

42 Varlamov, D.A., Kudin, A.P., Vielhaber, S., Schroder, R., Sassen, R., Becker, A., Kunz, D., Haug, K., Rebstock, J., Heils, A. et al. (2002) Metabolic consequences of a novel missense mutation of the mtDNA CO I gene. Hum Mol Genet, 11, 1797–1805.

43 Capaldi, R.A. (1990) Structure and function of cytochrome c oxidase. Annu Rev Biochem, 59, 569–596.

44 Sharma, L.K., Lu, J. and Bai, Y. (2009) Mitochondrial respiratory complex I: structure, function and implication in human diseases. Curr Med Chem, 16, 1266–1277.

45 Howell, N. (2003) LHON and other optic nerve atrophies: the mitochondrial connection. Dev Ophthalmol, 37, 94–108.

46 Alharbi, M.A., Al-Kafaji, G., Khalaf, N.B., Messaoudi, S.A., Taha, S., Daif, A. and Bakhiet, M. (2019) Four novel mutations in the mitochondrial ND4 gene of complex I in patients with multiple sclerosis. Biomed Rep, 11, 257–268.

47 Maassen, J.A., van Essen, E., van den Ouweland, J.M. and Lemkes, H.H. (2001) Molecular and clinical aspects of mitochondrial diabetes mellitus. Exp Clin Endocrinol Diabetes, 109, 127–134.

48 King, M.P., Koga, Y., Davidson, M. and Schon, E.A. (1992) Defects in mitochondrial protein synthesis and respiratory chain activity segregate with the tRNA(Leu(UUR)) mutation associated with mitochondrial myopathy, encephalopathy, lactic acidosis, and strokelike episodes. Mol Cell Biol, 12, 480–490.

49 van den Ouweland, J.M., Lemkes, H.H., Trembath, R.C., Ross, R., Velho, G., Cohen, D., Froguel, P. and Maassen, J.A. (1994) Maternally inherited diabetes and deafness is a distinct subtype of diabetes and associates with a single point mutation in the mitochondrial tRNA(Leu(UUR)) gene. Diabetes, 43, 746–751.

50 Chomyn, A., Martinuzzi, A., Yoneda, M., Daga, A., Hurko, O., Johns, D., Lai, S.T., Nonaka, I., Angelini, C. and Attardi, G. (1992) MELAS mutation in mtDNA binding site for transcription termination factor causes defects in protein synthesis and in respiration but no change in levels of upstream and downstream mature transcripts. Proc Natl Acad Sci U S A, 89, 4221–4225.

51 Goto, Y., Nonaka, I. and Horai, S. (1990) A mutation in the tRNA(Leu)(UUR) gene associated with the MELAS subgroup of mitochondrial encephalomyopathies. Nature, 348, 651–653.

52 Chen, F.L., Liu, Y., Song, X.Y., Hu, H.Y., Xu, H.B., Zhang, X.M., Shi, J.H., Hu, J., Shen, Y., Lu, B. et al. (2006) A novel mitochondrial DNA missense mutation at G3421A in a family with maternally inherited diabetes and deafness. Mutat Res, 602, 26–33.

53 Zhelankin, A.V. and Sazonova, M.A. (2012) [Association of the mutations in the human mitochondrial genome with chronic non-inflammatory diseases: type 2 diabetes, hypertension and different types of cardiomyopathy]. Patol Fiziol Eksp Ter, in press., 123–128.

54 Wortmann, S.B., Champion, M.P., van den Heuvel, L., Barth, H., Trutnau, B., Craig, K., Lammens, M., Schreuder, M.F., Taylor, R.W., Smeitink, J.A. et al. (2012) Mitochondrial DNA m.3242G > A mutation, an under diagnosed cause of hypertrophic cardiomyopathy and renal tubular dysfunction? Eur J Med Genet, 55, 552–556.

55 Ma, L., Wang, H., Chen, J., Jin, W., Liu, L., Ban, B., Shen, J., Hua, Z. and Chai, J. (2000) Mitochondrial gene variation in type 2 diabetes mellitus: detection of a novel mutation associated with maternally inherited diabetes in a Chinese family. Chin Med J (Engl), 113, 111–116.

56 Ohkubo, E., Aida, K., Chen, J., Hayashi, J.I., Isobe, K., Tawata, M. and Onaya, T. (2000) A patient with type 2 diabetes mellitus associated with mutations in calcium sensing receptor gene and mitochondrial DNA. Biochem Biophys Res Commun, 278, 808–813.

